# Home collection of nasal swabs for detection of influenza in the Household Influenza Vaccine Evaluation Study

**DOI:** 10.1101/2020.03.24.20042556

**Authors:** Ryan E. Malosh, Joshua G. Petrie, Amy P. Callear, Arnold S. Monto, Emily T. Martin

## Abstract

**Background:** Community based studies of influenza and other respiratory viruses (e.g. SARS-COV-2) require laboratory confirmation of infection. During the current COVID-19 pandemic, social distancing guidelines require alternative data collection in order protect both research staff and participants.

Home-collected respiratory specimens are less resource intensive, can be collected earlier after symptom onset, and provide a low-contact means of data collection. A prospective, multi-year, community-based cohort study is an ideal setting to examine the utility of home-collected specimens for identification of influenza.

**Methods:** We describe the feasibility and reliability of home-collected specimens for the detection of influenza. We collected data and specimens between October 2014 and June 2017 from the Household Influenza Vaccine Evaluation (HIVE) Study. Cohort participants were asked to collect a nasal swab at home upon onset of acute respiratory illness. Research staff also collected nose and throat swab specimens in the study clinic within 7 days of onset. We estimated agreement using Cohen’s kappa and calculated sensitivity and specificity of home-collected compared to staff-collected specimens.

**Results:** We tested 336 paired staff- and home-collected respiratory specimens for influenza by RT-PCR; 150 staff-collected specimens were positive for influenza A/H3N2, 23 for influenza A/H1N1, 14 for influenza B/Victoria, and 31 for influenza B/Yamagata. We found moderate agreement between collection methods for influenza A/H3N2 (0.70) and B/Yamagata (0.69) and high agreement for influenza A/H1N1 (0.87) and B/Victoria (0.86). Sensitivity ranged from 78-86% for all influenza types and subtypes. Specificity was high for influenza A/H1N1 and both influenza B lineages with a range from 96-100%, and slightly lower for A/H3N2 infections (88%).

**Conclusions:** Collection of nasal swab specimens at home is both feasible and reliable for identification of influenza virus infections.

## Background

Influenza is a respiratory virus that causes substantial annual morbidity and mortality, including an estimated 200,000 hospitalizations and 30,000 deaths in the United States each year [1]. Vaccines are available for prevention of influenza virus infections, but recent estimates have shown only moderate vaccine effectiveness (VE) [2,3]. Annual variation is common, in terms of the frequency and severity of infection as well as in VE. As a result, further studies of influenza VE and transmission are needed with the goal of improving control.

Prospective, longitudinal community-based studies have a broad range of applications in respiratory virus epidemiology. These studies will be essential to better understand the extent of the pandemic caused by the novel coronavirus SARS-COV-2 and the full range of COVID-19 illness. These studies also present unique opportunities to explore more in-depth questions about immune correlates of influenza vaccine failure as well as susceptibility to and transmission of infection. Nevertheless, they are much more resource intensive than comparably sized studies using case-control designs [4].

Ensuring adequate and timely specimen collection across a large cohort is particularly important as the circulation of respiratory viruses varies greatly on both a seasonal and annual basis. The minimum detectable effect size for preventive interventions in these studies is particularly sensitive to variations in the infection risk. Sensitive methods of pathogen detection (e.g. RT-PCR) have improved identification of cases, but specimen collection methods that are timely and reduce the burden on study participants are needed to minimize the likelihood of incorrectly determining infection status [5].

There have been several feasibility and validation studies which have suggested that self- or parent-collected nasal swabs are both acceptable and result in quality specimens for identification of respiratory viruses [5–11]. There are few studies, however, involving community-based participants collecting specimens in their own homes, outside of a study clinic setting [12]. We sought to describe the feasibility of home-collected respiratory specimens collected at home during an acute respiratory infection (ARI) and examine the validity of the specimens compared to those collected by research staff.

## Methods

We collected respiratory specimens at two time points during ARI to examine the feasibility and reliability of home-collected specimens in three seasons (2014-2015 through 2016-2017) of the Household Influenza Vaccine Evaluation (HIVE) Study. HIVE, a prospective cohort study of influenza and other respiratory viruses in households with children, has been ongoing since 2010. Eligible households were those with at least 3 individuals, at least two of whom were children < 18 years old. At enrollment, participants completed demographic and health history questionnaires and were provided with instructions on collection of nasal swab specimens at home on the first day of an eligible ARI. This study was reviewed and approved by the institutional review board at the University of Michigan Medical School.

### ARI Surveillance

Active surveillance for identification of ARI was conducted year round, beginning in October 2014. Participating households were instructed to contact the study team at the onset of new ARI and were additionally contacted each week by email or telephone. Report of an illness meeting the study case definition triggered collection of an upper respiratory specimens at home on the first day of symptoms (home-collected) and then in the study clinic within 7 days of symptom onset (staff-collected).

### Home-collected respiratory specimens

Participants (or their parents) were asked to collect a nasal swab specimen at home on the first day of their illness (home-collected specimens). Adults were trained in person to collect specimens prior to illness season at enrollment visits. In addition, each household was given an instruction card and a link to an online video with detailed instructions on how to collect nasal swab specimens. Home-collected specimens were stored in commercially prepared viral transport media and submitted to research staff during their scheduled illness visit in the study clinic.

### Staff-collected respiratory specimens

Research staff scheduled a specimen collection visit at the study clinic within 7 days of illness onset. At these visits, oropharyngeal and mid-turbinate swabs (mid-turbinate only in children <3 years of age) were collected by research staff and combined in commercially prepared viral transport media (staff-collected specimens). Specimens were kept at room temperature until they were transported to laboratory.

### Influenza Testing

Specimens were tested by RT-PCR for laboratory confirmation of influenza, using primers and probes from the Centers for Disease Control and Prevention. Influenza subtype was determined for influenza A positive specimens and lineage was determined for influenza B positive specimens.

Specimens were also tested for human ribonuclease P (RNP), using a Cycle threshold (Ct) cutoff of ≤40 to determine specimen quality. [13,14] Specimens without detection of RNAseP were excluded from further analyses.

All staff-collected specimens were tested for influenza by RT-PCR. We selected a subset of paired home-collected specimens for influenza testing if staff-collected specimen was RT-PCR confirmed influenza positive, if staff-collected specimen test results were inconclusive for subtype, or if the onset of symptoms was within 7 days of an influenza case in a household contact. In all cases the specimen collected by staff was within 7 days of onset.

### Statistical analysis

We first described the proportion of ARI with home-collection of specimens by season, and participant and household characteristics. We examined the timing of home-collection and clinic collection in relation to the onset of symptoms. Finally, we examined the reliability of home- and staff-collected specimens. Dichotomous outcomes, including detection of influenza, were compared by calculating Cohen’s kappa. We interpreted the kappa statistic as suggested by McHugh et al: 0-0.20 no agreement, 0.21-0.39, minimal; 0.40-0.59, weak; 0.60-0.79 moderate; 0.80-0.90 strong; and > 0.90 almost perfect [15]. We also examined the sensitivity and specificity of home-collected specimens using staff-collected specimens as the gold standard. Mean Cycle threshold (Ct) values among concordant influenza positive specimens were compared using the paired Wilcoxon test.

## Results

### Study Population

We followed 1431 individuals during the 2014-15 season, 996 individuals in 2015-16 and 890 in 2016-17. In each season, the study population included approximately 60% children < 18 years and approximately 50% female participants (Table 1). In 2014-15, 702 individuals reported 1362 ARI and had respiratory specimens collected by study staff, 452 individuals reported 934 ARI in 2015-16, and 416 reported 810 ARI, in 2016-17. In all study years, children < 5 years old and female participants were over-represented among ARI events compared to the overall study population (Table 1). Approximately 70-80% of ARI with staff-collection of respiratory specimens in each year also included home-collection of nasal swabs. The subset of 336 paired home- and staff-collected specimens that were selected for influenza testing were representative of all ARI with specimen collection (Table 1).

**Table 1.** Individual and household characteristics for the study population overall and among all ARI with staff-collected specimens, all ARI with home-collected specimens and paired staff- and home-collected specimens tested for influenza, by season.

|  | Study Population<br>n (%) <sup>1</sup> | ARI with specimen collection<br>n (%) <sup>1</sup> |  | Influenza<br>testing subset,<br>n (%) <sup>1</sup> |
| --- | --- | --- | --- | --- |
| Season |  | Staff collected | Home collected |  |
| **2014-2015** | **1431** | **1362** | **997** | **214 (21) <sup>2</sup>** |
| Age group |  |  |  |  |
| < 5 years | 200 (14) | 306 (22) | 215 (22) | 38 (18) |
| 5-11 years | 442 (31) | 414 (30) | 328 (33) | 83 (39) |
| 12-17 years | 220 (15) | 153 (11) | 110 (11) | 22 (10) |
| 18+ years | 569 (40) | 489 (36) | 344 (34) | 71 (33) |
| Sex |  |  |  |  |
| Female | 746 (52) | 785 (58) | 576 (58) | 115 (54) |
| Male | 685 (48) | 577 (42) | 421 (42) | 99 (46) |
| **2015-2016** | **996** | **934** | **700** | **39 (6) <sup>2</sup>** |
| < 5 years | 145 (15) | 216 (23) | 168 (24) | 8 (21) |
| 5-11 years | 317 (32) | 262 (28) | 205 (29) | 17 (44) |
| 12-17 years | 141 (14) | 108 (12) | 73 (10) | 0 (0) |
| 18+ years | 393 (39) | 348 (37) | 254 (36) | 14 (36) |
| Sex |  |  |  |  |
| Female | 520 | 524 | 404 | 23 (59) |
| Male | 476 | 410 | 296 | 16 (41) |
| **2016-2017** | **890** | **810** | **642** | **83 (13) <sup>2</sup>** |
| < 5 years | 113 (13) | 162 (20) | 129 (20) | 9 (11) |
| 5-11 years | 261 (29) | 252 (31) | 203 (32) | 31 (37) |
| 12-17 years | 148 (17) | 99 (12) | 81 (13) | 17 (20) |
| 18+ years | 368 (41) | 294 (36) | 226 (35) | 26 (31) |
| Sex |  |  |  |  |
| Female | 457 (51) | 424 (52) | 345 (54) | 45 (54) |
| Male | 433 (49) | 383 (47) | 294 (46) | 38 (46) |

### Timing of collection

Among the paired specimens tested for influenza, 313 (93%) were collected within 2 days of illness onset. The median duration from onset to home-collection of respiratory specimens was 0 days (IQR 0-1) compared to 2 days (IQR 1-4) for staff-collected specimens (Figure 1). Sixty-seven (20%) home-collected specimens were collected on the same day as staff-collected specimens. 109 (32%) home-collected specimens were collected 1 day prior to staff-collected specimens. The remaining 160 (48%) were collected ≥2 days prior to staff-collected specimens. Human RNase-P gene was detected at a Ct of less than 40 in all tested home-collected specimens and in all but one staff-collected specimens. The staff-collected specimen with RNase-P Ct>40 was excluded from further analysis.

**Figure 1.**
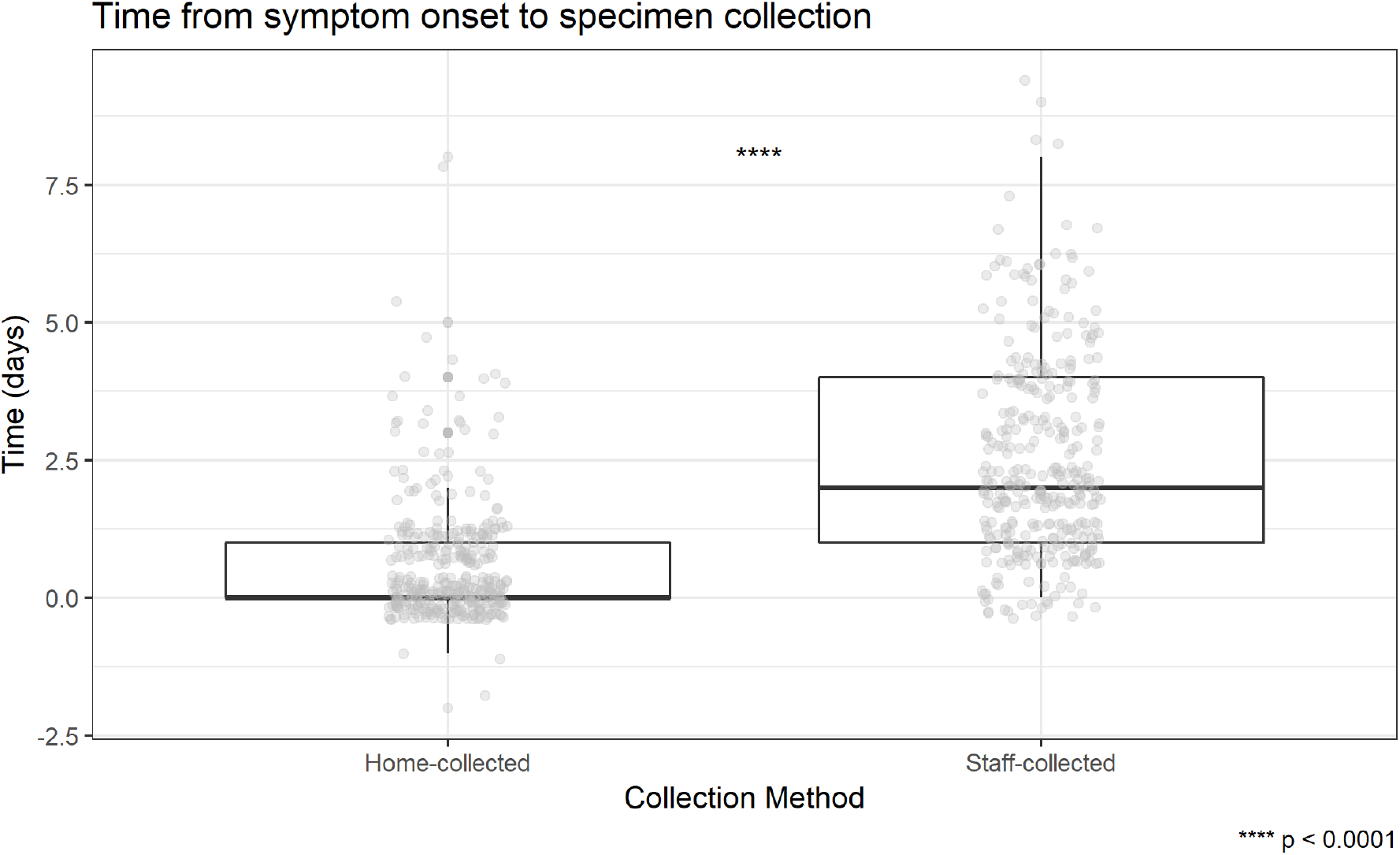
Time (days) from symptom onset to specimen collection for paired home-collected and staff-collected respiratory specimens.

### Influenza testing

We identified 150 cases of influenza A/H3N2 infection in staff-collected specimens. 123 of these infections were confirmed by testing home-collected specimens. An additional 22 cases of influenza A/H3N2 infection were identified by testing home-collected specimens (Table 2). Similarly, we identified 32 cases of influenza B/Yamagata virus infection in staff-collected specimens. 25 of these were confirmed and an additional 12 cases were identified by testing home-collected specimens. We also identified 23 cases of influenza A/H1N1 and 14 cases of B Victoria (Table 2).

**Table 2.**
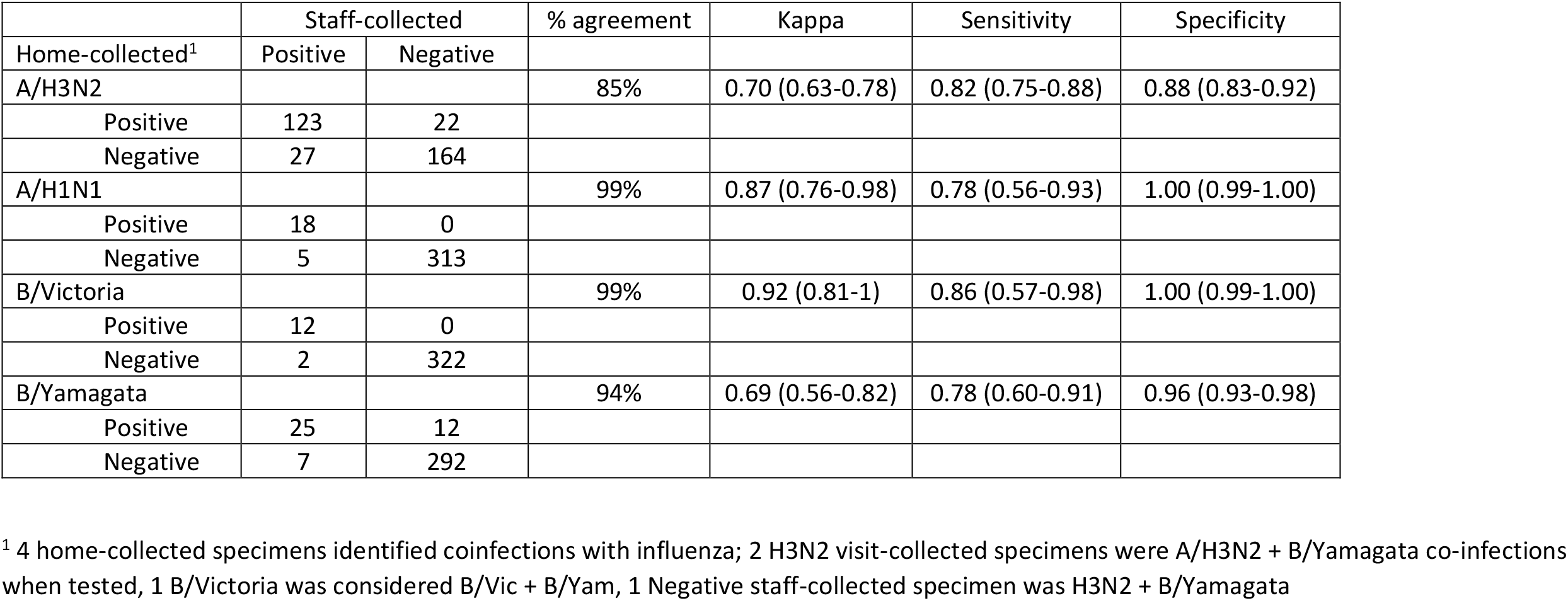
Influenza testing results in paired home- and staff-collected specimens, by influenza type and subtype.

The agreement between home- and staff-collected specimens was 85% for influenza A/H3N2 viruses, but increased to 99% when analysis was restricted to paired specimens collected within 1 day of each other. The other influenza viruses we tested for also had extremely high agreement inclusive of all collection times: 99% for influenza A/H1N1, 99% for influenza B/Victoria, and 94% for influenza B/Yamagata. We used Cohen’s kappa to estimate agreement (0-0.20, no agreement, 0.21-0.39, minimal; 0.40-0.59, weak; 0.60-0.79 moderate; 0.80-0.90 strong; and > 0.90 almost perfect agreement.) We found moderate agreement between home- and staff-collected specimens for influenza A/H3N2 and B/Yamagata viruses, strong agreement for influenza A/H1N1, and almost perfect agreement for B/Victoria viruses (Table 2). Sensitivity was reasonably high for all influenza types and subtypes, ranging from 78-86%. Specificity was extremely high for influenza A/H1N1 and both influenza B lineages with a range from 96-100%, and slightly lower for A/H3N2 infections (88%).

### RT-PCR Cycle Threshold (Ct) Values

Overall, the median Ct for staff-collected specimens was 24.4 (IQR 21.60-31.19) for influenza B/Victoria, 26.58 (IQR 23.08-31.56) for influenza A/H3N2, 27.18 (IQR 23.86-31.44) for influenza B/Yamagata, and 30.93 (IQR 24.55-34.58) for influenza A/H1N1pdm09. These values were not significantly different (p = 0.1661). We also compared the mean Ct values for paired specimens that had concordant influenza testing results, stratified by the time between home- and staff-collected specimens (Figure 2). For influenza A, we found that home-collected specimens had a lower median Ct value, which is generally consistent with a higher viral load, (22.77, IQR 19.88-27.91) than staff-collected specimens (28.93, IQR 24.51-31.5) when collected ≥ 2 days prior to staff-collected specimens (p<0.001). For influenza B viruses we found no significant differences in Ct value after stratifying on time between home- and staff-collected specimens (Figure 2). Thirty-eight staff- and home-collected specimens were collected on the same day. Among these, median Ct values were slightly lower for home-collected specimens (23.68 IQR 20.51-28.62) compared to staff-collected specimens (26.07 IQR 23.08-31.56) for influenza A/H3N2 viruses (p = 0.004). No differences in median Ct values were observed among the other influenza subtypes.

**Figure 2.**
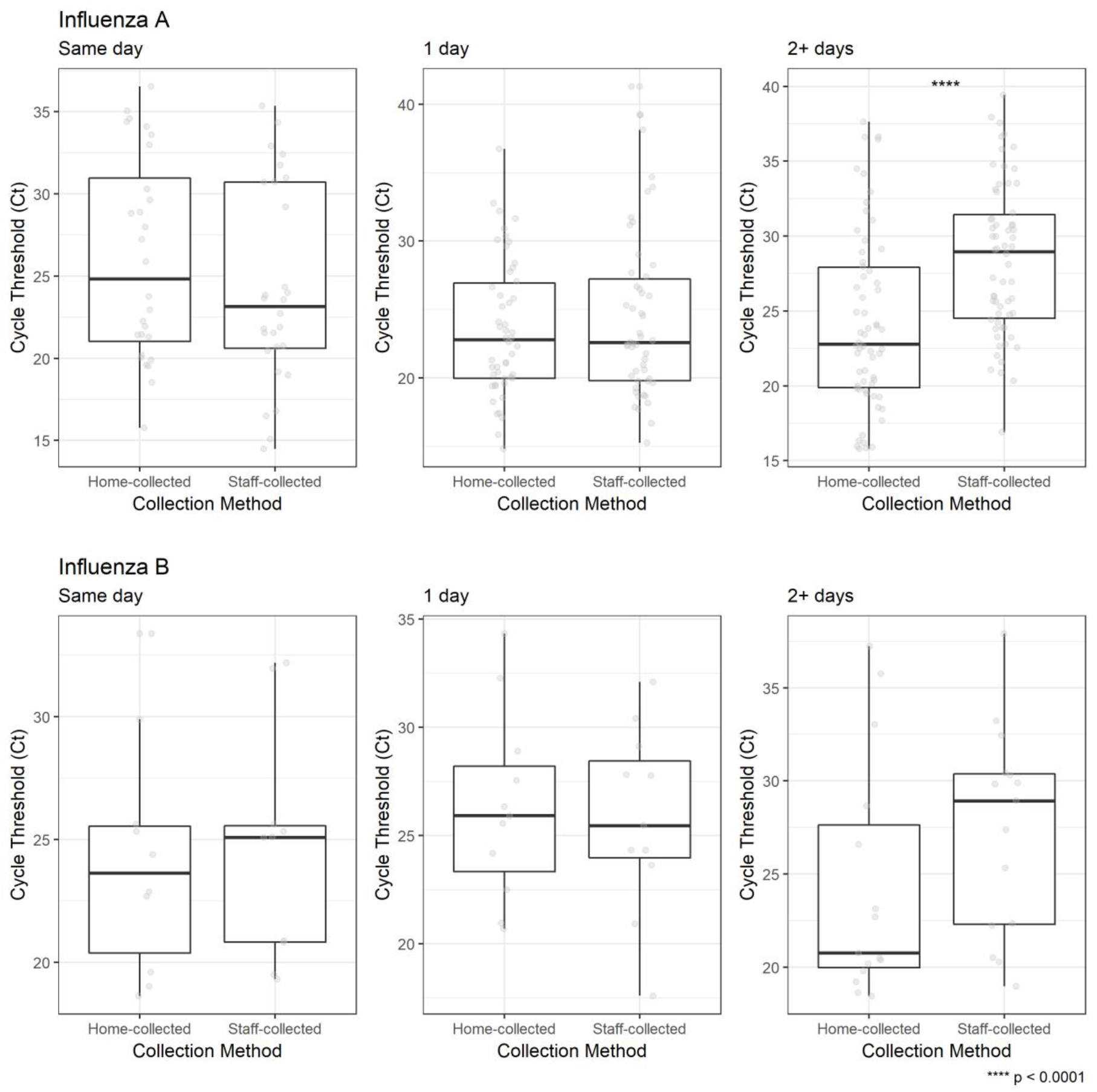
Ct Value for home-collected and staff-collected respiratory specimens by influenza type, stratified by time (days) between home- and staff-collection.

We also compared Ct values between home- and staff-collected specimens by vaccination status for each influenza type/subtype. Median Ct value was lower for home-collected specimens compared to staff-collected specimens among influenza A/H3N2 positive specimens for both vaccinated (23.57 vs 26.42, p=0.001) and unvaccinated individuals 22.31 vs 27.13, p=0.011). We found no difference in Ct value comparing vaccinated to unvaccinated influenza A/H3N2 cases for either home-collected (23.57 vs 22.31, p=0.543) or staff-collected specimens (26.42 vs 27.13, p=0.705).

## Discussion

We found that unsupervised, home-collection of respiratory specimens for identification of influenza infection was both feasible and reliable. A high proportion of participants completed the home collection and the vast majority completed collection within 2 days of symptom onset. Overall, the percent agreement was high for all influenza subtypes examined in this study. Agreement between home and staff-collection was moderate for influenza A/H3N2 and B/Yamagata viruses and higher for A/H1N1 and B/Victoria viruses. Additionally, assessment of RNAseP detection suggested that specimen quality was adequate for home-collected specimens.

Previous studies have demonstrated that nasal swabs collected by research participants under the supervision of research or clinical staff are feasible for the identification of respiratory virus infections [6–9,12,16,17]. In many of these studies acceptability of self- or parent-collection was preferred to investigator or clinician collection [7,16]. In the few studies of unsupervised collection, self-collected specimens were collected and returned in the timeframe recommended by the research team [6,16]. In addition, specimen quality in many studies has been similar regardless of collection method [5,19]. A Canadian study found that both viral loads and RNaseP were similar between self-collected and investigator-collected specimens [8]. A study in pregnant women found 100% of self-collected specimens detected RNaseP [6], as we found in the current study. Other measures of specimen quality have also been similar between collection methods [11,18]. Importantly, many of these feasibility studies were pilot studies with relatively small sample sizes, and self-collection was often completed in the presence of the study investigators. Our study confirms the feasibility and timeliness of unsupervised collection of respiratory specimens in a large, longitudinal cohort study. As we are currently experiencing with COVID-19 pandemic, unsupervised collection Is essential to provide maximum protection to research staff and participants and to comply with social distancing guidelines from public health authorities.

High levels of agreement and high sensitivity and specificity of self-collected specimens have also been demonstrated in a variety of settings. For example, self-collected nasal swabs have been compared to nasal wash with high levels of sensitivity (88-95%) [9]. A study of children < 5 years comparing supervised parental collection to pediatrician collected specimens found similarly sensitivity (89%) and high specificity (97%) [7]. Additional studies have established that self-collected specimens are adequately sensitive and specific for detection of respiratory viruses [8,17,19]. We observed similarly high reliability of home-collected specimens in terms of sensitivity and specificity.

The overall agreement was lower in our study compared to others [7,8,11] for influenza A/H3N2 viruses in part because we identified 29 additional influenza infections in participants who were influenza negative according to their staff-collected specimens. The difference in agreement with previous studies may be explained by the timing of our collection methods. Many of the previous studies involved concurrent collection of self- and staff-collected specimens, whereas our specimens are collected on onset (home-collected) and again within 7 days of onset (staff-collected). One study of parent-collected specimens found that time from symptom onset to collection was the only factor associated with respiratory virus positivity; collection method and subjective quality of parent-collected specimens were not associated with detection [6]. Likewise, we found that time between home- and staff-collection was an important factor in terms of both Ct value and agreement. Agreement in our study was higher when analyses were restricted to paired specimens collected within 1 day of each other. Our coupling of home-collection with prospective, active surveillance has allowed us to capture viral specimens on the day of onset without the delay inherent in scheduling a visit with research staff. Future applications of this method can include sequential specimen collection for studies of viral shedding over time and virus transmission.

In conclusion, in a large community-based study, we have demonstrated that collection of nasal swab specimens at home is both feasible and reliable for identification of respiratory virus infections. We also demonstrate that there is added utility to self-collection of nasal swab specimens earlier in an ARI episode, specifically by identifying additional infections. Home-collected respiratory specimens can be used in large, community-based observational studies of respiratory viruses to facilitate identification of laboratory confirmed infections.

## Data Availability

The data that support the findings of this study are available from the corresponding author, REM, upon reasonable request.

## Acknowledgements

The authors wish to thank the HIVE Study participants for their participation in this research, as well as the HIVE study staff for their hard work and dedication to the project.

